# The impact of perioperative diagnostic tools on clinical outcomes and cost-effectiveness in parathyroid surgery: a health economic evaluation

**DOI:** 10.1101/2023.12.04.23299113

**Authors:** Daniel Bátora, Rowan Iskandar, Jürg Gertsch, Reto M. Kaderli

**Affiliations:** Institute of Biochemistry and Molecular Medicine, University of Bern, Bern, Switzerland; Graduate School of Cellular and Biomedical Sciences, University of Bern, Bern, Switzerland; Center of Excellence in Decision-Analytic Modeling and Health Economics Research, sitem-insel, Bern, Switzerland; Department of Health Services, Policy, & Practice, Brown University, Providence, RI, USA; Institute of Social and Preventive Medicine, University of Bern, Bern, Switzerland; Multidisciplinary Center for Infectious Diseases, University of Bern, Bern, Switzerland; Department of Visceral Surgery and Medicine, Inselspital, Bern University Hospital, University of Bern, Bern, Switzerland

**Keywords:** cost-effectiveness, parathyroid surgery, preoperative imaging, primary hyperparathyroidism

## Abstract

**Objectives:** Pre- and intraoperative diagnostic tools influence the surgical management of primary hyperparathyroidism (PHPT), whereby their performance of classification varies considerably for the two common causes of PHPT: solitary adenomas and multiglandular disease. A consensus on the use of such diagnostic tools for optimal perioperative management of all PHPT patients has not been reached.

**Design:** A decision tree model was constructed to estimate and compare the clinical outcomes and the cost-effectiveness of preoperative imaging modalities and intraoperative parathyroid hormone (ioPTH) monitoring criteria in a 14-year time horizon. The robustness of the model was assessed by conducting a one-way sensitivity analysis and probabilistic uncertainty analysis.

**Setting:** The United States healthcare system.

**Population:** A hypothetical population consisting of 5,000 patients with sporadic, symptomatic, or asymptomatic PHPT.

**Interventions:** Pre- and intraoperative diagnostic modalities for parathyroidectomy

**Main outcome measures:** Costs, quality-adjusted life years (QALYs), net monetary benefits (NMB), clinical outcomes

**Results:** In the base-case analysis, four-dimensional (4D)-computed tomography (CT) was the least expensive strategy with $10,289 and 13.93 QALYs. Ultrasound and ^99m^Tc-Sestamibi single-photon-emission computed tomography/CT were both dominated strategies, while ^18^F-fluorocholine positron emission tomography was cost-effective with a net monetary benefit of $264 considering a willingness to pay threshold of $95,958. The application of ioPTH monitoring with the Vienna criterion decreased the rate of reoperations from 10.50 to 0.58 per 1,000 patients. Due to an increased rate of bilateral neck explorations from 257.45 to 347.45 per 1,000 patients, it was not cost-effective.

**Conclusions:** 4D-CT is the most cost-effective instrument for the preoperative localization of parathyroid adenomas. Due to an excessive increase of bilateral neck explorations, the use of ioPTH monitoring is not cost-effective in PHPT but leads to a significant reduction of reoperations.

**Strengths and limitations of the study:**

- Our decision tree model is the most complete for parathyroidectomy; incorporating both solitary adenomas and multiglandular disease and ioPTH monitoring.
- In addition to cost-effectiveness, we present the impact of the interventions on the major clinical outcomes.
- Our study is limited to the United States and does not include a societal perspective.
- The model did not consider the potential institutional variations in the prevelance of multiglandular disease
- There remains uncertainty for certain parameters for the model as they were derived from a limited number of single-institution studies

## 1. INTRODUCTION

Primary hyperparathyroidism (PHPT) is a common endocrine disorder with a prevalence of one to seven cases per 1,000 adults, and is the primary reason for hypercalcemia.^1^ In 70 - 90%, 10 – 30%, and less than 1% of the cases, the underlying cause is a single gland parathyroid adenoma, multiglandular disease (MGD), and parathyroid carcinoma, respectively.^2^ Even though PHPT is often diagnosed at an asymptomatic stage, the surgical removal of the diseased tissue is generally recommended due to the long-term deleterious effects of PHPT and remains the only curative treatment.^3^

To optimize surgical cure rates and to reduce operative trauma, surgeons often rely on imaging technologies and specialists for the preoperative localization of the diseased gland(s). If a solitary adenoma is suspected, a focused parathyroidectomy (FP) might be performed with a cure rate that is comparable to the conventional bilateral neck exploration (BNE).^4^ FP is associated with reduced operative times, risk of developing postoperative hypoparathyroidism and recurrent laryngeal nerve (RLN) injury.^5^ However, the sensitivity and specificity of a preoperative localization vary not only between the different preoperative diagnostic modalities but also between single adenomas and MGD.^6–9^

If an FP is performed, an intraoperative parathyroid hormone (ioPTH) monitoring is recommended to exclude MGD and to avoid reoperations.^10^ The two most common criteria for defining surgical success, measured as a 50% decline of the ioPTH level 10 minutes after resection, use different baselines. The Miami criterion uses the highest ioPTH level (pre-incision or pre-excision) as the baseline, while the Vienna criterion uses the pre-incision ioPTH level as the baseline.^11^

The overall clinical utility of ioPTH monitoring is under debate due to the risk of misleading surgeons with false negatives and false positive results depending on the criterion used.^12^

Furthermore, the application of ioPTH monitoring would require additional resources: increased surgery time and the cost of the procedure.^13^ Some recent cost-effectiveness analyses focused only on preoperative imaging modalities without considering the inclusion of patients with MGD and reoperations.^14^ Other health economic evaluations only assessed the cost of preoperative localization and ioPTH monitoring, omitting the clinical outcomes.^12^ The absence of a complete evidence on the clinical benefits and cost-effectiveness of using either criterion for the ioPTH evaluation contributes to the lack of consensus on whether the use of ioPTH monitoring is warranted and what criterion should be for solitary adenomas and MGD.^12^

Thus, this study aims to estimate the clinical outcomes and the cost-effectiveness of common preoperative imaging modalities and ioPTH monitoring criteria for patients with sporadic, symptomatic, or asymptomatic PHPT.

## 2. METHODS

### 2.1 Decision-analytic model

We structured the clinical decision-making problem using the decision-analytic modeling framework, where the architecture of the model is in the form of a decision-tree model. We simulated a hypothetical population consisting of 5,000 65-year-old patients with sporadic, symptomatic, or asymptomatic PHPT who met the recently updated National Institutes of Health (NIH) criteria for parathyroid surgery and were eligible for Medicare reimbursements.^10^ The available strategies included four preoperative imaging modalities: ultrasound, ^99m^Tc-Sestamibi single-photon-emission computed tomography (SPECT)/computed tomography (CT), four-dimensional (4D)-CT, ^18^F-fluorocholine positron emission tomography (FCH-PET/CT). For each preoperative imaging modality, the use of ioPTH monitoring was considered. The baseline value below which a decrease of ioPTH concentration is considered positive was determined by applying either the Miami or Vienna criterion.

For patients with a solitary adenoma, an FP was performed after a successful preoperative localization and an adequate decrease of ioPTH. A false negative localization of a solitary adenoma resulted in BNE, while a false positive localization led first to a unilateral neck exploration (UNE) and in cases where the adenoma was located on the contralateral side to a BNE.

For patients with MGD, a true positive localization and a true positive decrease of ioPTH led to either a unilateral neck exploration (UNE) or BNE, depending on the localization of the adenomas, whereas a false negative decrease of ioPTH resulted in a BNE. A false positive localization of MGD with a false positive decrease of ioPTH leads to a reoperation after UNE or BNE. A false positive localization of MGD with a true negative decrease of ioPTH leads to a BNE. A false negative localization leads to a BNE. Fig 1 depicts a simplified version of the decision-tree model.

**Figure 1.**
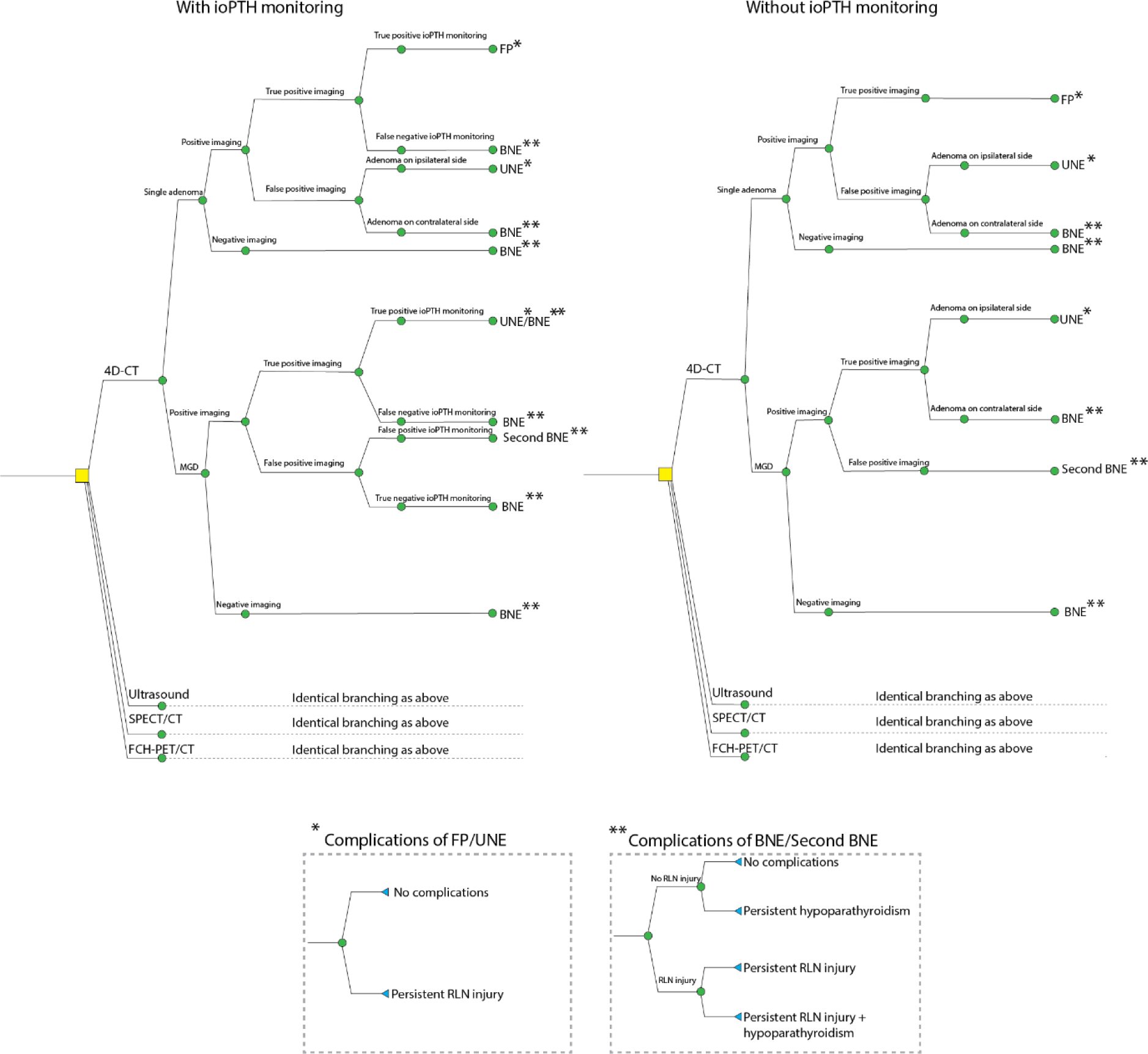
Simplified illustration of the decision-tree model. Patients diagnosed with primary hyperparathyroidism (PHPT) due to a single adenoma or multiglandular disease (MGD) had a preoperative imaging (four-dimensional [4D]-computed tomography [CT], ultrasound, ^99m^Tc-Sestamibi single-photon-emission computed tomography [SPECT]/CT or ^18^F-fluorocholine positron emission tomography [FCH-PET/CT]). The use of intraoperative parathyroid hormone (ioPTH) monitoring was considered for each intervention. Based on the result of preoperative imaging and ioPTH monitoring, either focused parathyroidectomy (FP), unilateral neck exploration (UNE) or bilateral neck exploration (BNE) was performed. The following surgical complications were considered: persistent hypoparathyroidism and persistent recurrent laryngeal nerve (RLN) injury.

We incorporated the following assumptions into the model. First, an adenoma can always be differentiated macroscopically from a normal parathyroid gland by the surgeon in the absence of ioPTH monitoring. Second, in case of a successful preoperative localization with an adequate decrease of ioPTH, the surgeon would base his decisions solely on the ioPTH decrease and refrain from further explorations. This assumption implies that, in the case of a false positive localization of a single adenoma, the surgeon would perform a UNE/BNE irrespective of the ioPTH readout. Therefore, we did not consider false positive and true negative ioPTH readouts for these events. Since a false positive localization misses an adenoma in MGD, we considered false positive and true negative ioPTH readouts in this case. Third, the time horizon of the analysis was the life expectancy of a 65-year-old patient, which was estimated to be 14 years based on the US life table^15^ and was assumed to be uniform across the different surgeries. Based on expert advice, we assumed that the probability of complications, costs, and utilities for UNE are identical to those of FP. For reoperations, we considered the same model architecture with an increased risk of complications.^16^ Lastly, we assumed FP and BNE to be carried out in an outpatient and inpatient setting, respectively.

When available, estimates for the probability parameters were derived from the latest systematic reviews. The prevalence of solitary adenomas was taken from the latest surgical guideline by the American Association of Endocrine Surgeons, which was based on a comprehensive review of published papers from 1985 to 2015 by a multidisciplinary panel of experts.^17^ The sensitivity and positive predictive value (PPV) of ultrasonography in MGD were obtained from a systematic literature review between 1995 and 2003.^7^ The sensitivity and PPV of SPECT/CT, 4D-CT and FCH-PET/CT were retrieved from the latest systematic reviews ^6,8^, whereas the sensitivity and PPV for FCH-PET/CT in MGD was taken from a single-institution study.^9^ The sensitivity and specificity of the different ioPTH criteria were estimated from two single-institution studies with 570 patients.^11,18^ The probabilities of developing persistent postoperative hypoparathyroidism and persistent RLN injury were estimated from a randomized controlled trial with 91 patients.^19^ Table 1 lists all the model parameters’ estimates and their sources.

**Table 1.**
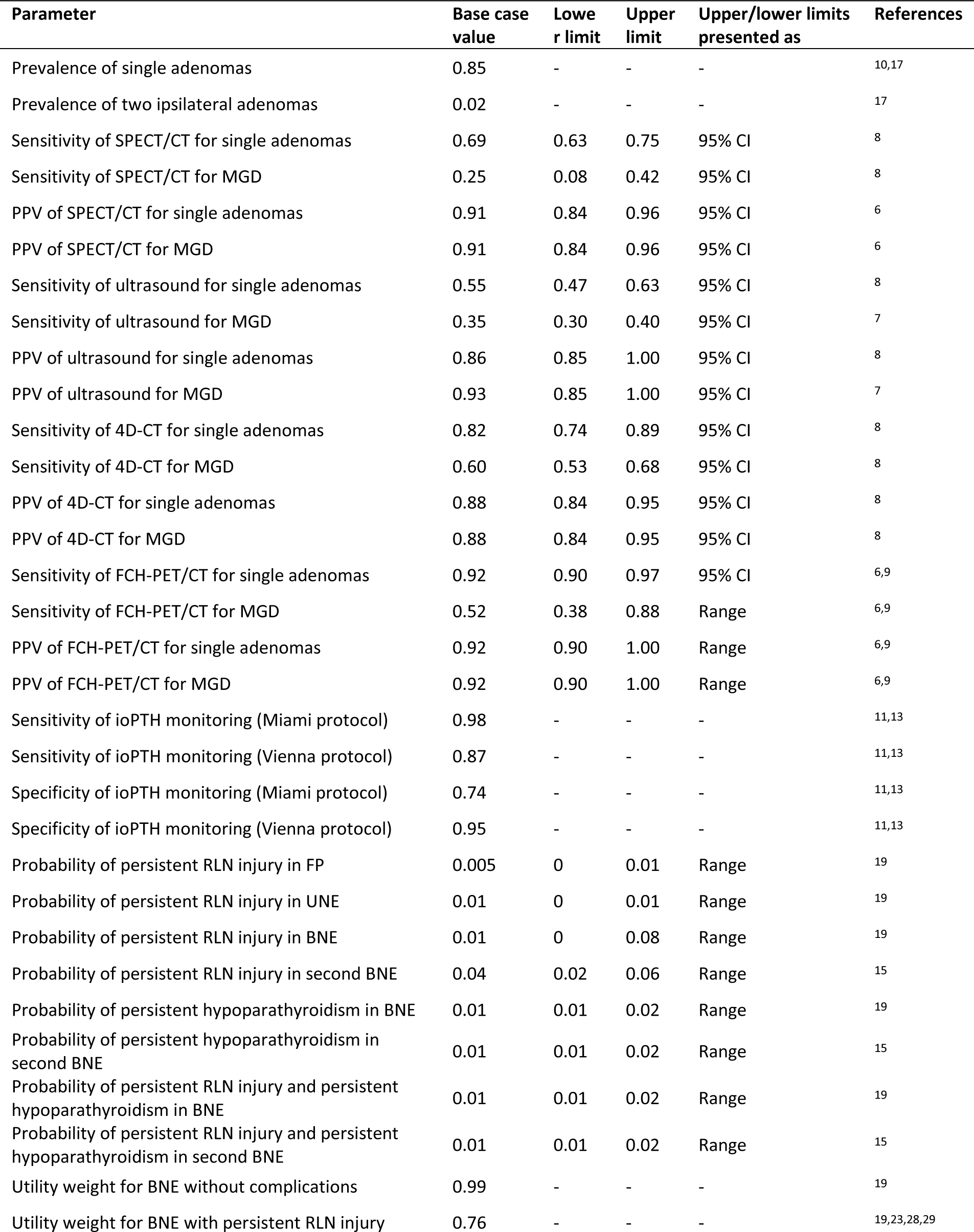
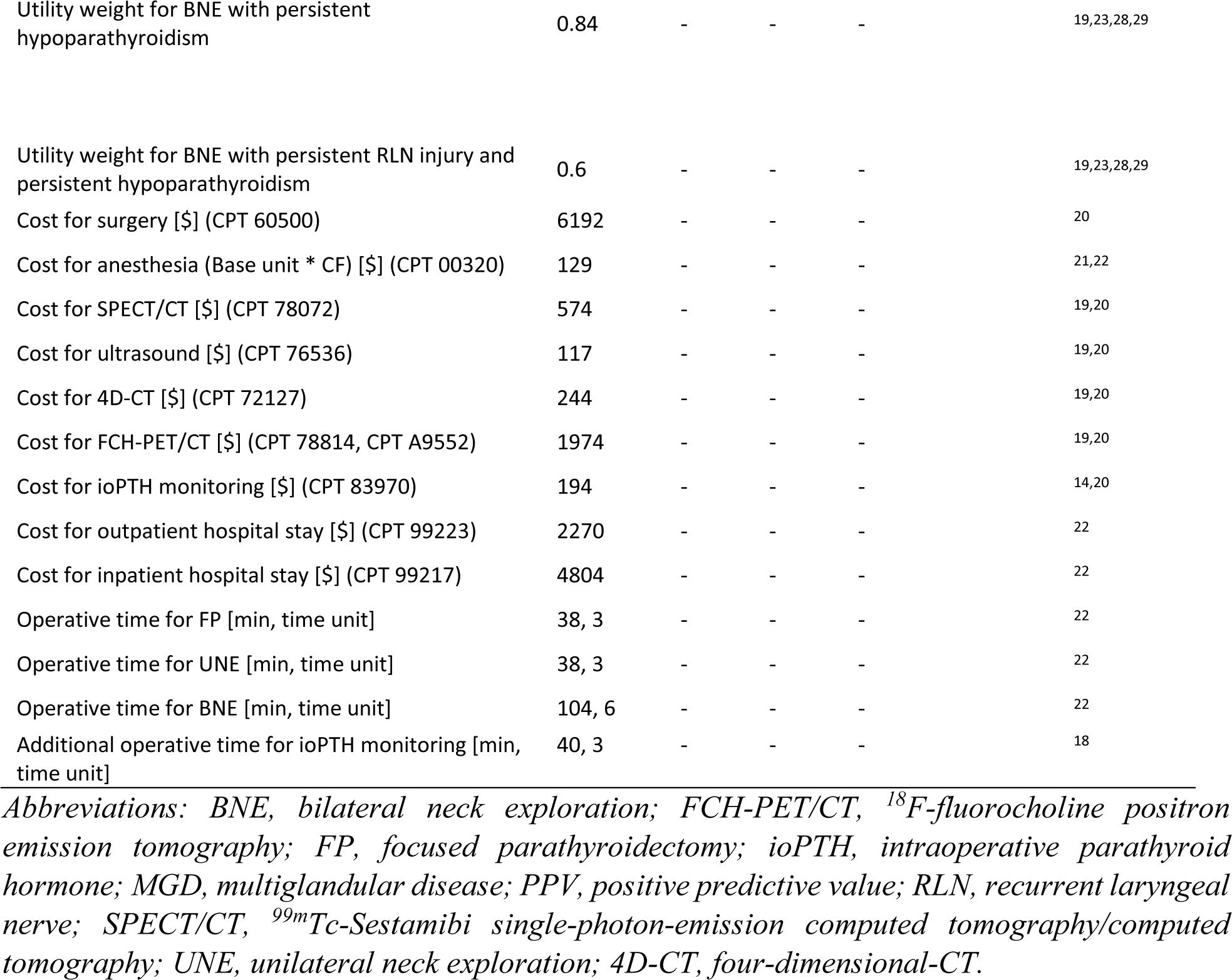
Estimated values for each parameter in the decision-tree model.

### 2.2 Cost-effectiveness analysis

The cost-effectiveness analysis was conducted from the healthcare system perspective, focusing on the United States. To calculate the total costs for each strategy, we considered the costs of the preoperative imaging modality, ioPTH assay, hospital stay (depending on the procedures), and the surgery time multiplied by the costs of the operating theater per minute. Costs were reported in 2022 US dollars. Current Procedural Terminology (CPT)-based physician and facility costs of surgery (CPT 60500), imaging (CPT 78072, CPT 76536, CPT 72127, CPT 78814, CPT A9552), and ioPTH monitoring (CPT 83970) were derived from the US medical reimbursement schedule.^20^ As FCH-PET/CT is currently not authorized in the United States, we combined the cost of a neck PET/CT scan (CPT 78814) with the price of the most commonly used radiotracer, ^18^F-fluorodeoxyglucose (CPT A9552).

We calculated anesthesiology fees (CPT 00320) by multiplying the base and time units, and the national average anesthesia conversion factor (CF) taken from the 2022 Centers for Medicare & Medicaid Services (CMS) release.^21^ The base unit for CPT 00320 was 6, and the time units were expressed in 15-minute increments. The national average anesthesia CF was 21.5623. The length of surgeries was converted into time units. The costs for outpatient (CPT 99223) and inpatient (CPT 99217) hospital services were derived from a previous study ^22^, which calculated the costs for the Diagnostic-Related Group (DRG) 627.

We expressed the effectiveness of each intervention as quality-adjusted life years (QALYs), which were calculated by multiplying the life expectancy by utility weights. The utility weights for persistent hypoparathyroidism and RLN injury, where a utility value of 1 corresponds to a perfect state of health and 0 corresponds to being dead, were derived from previous studies that used Short Form 36-Item Health Survey.^23^ As a metric for cost-effectiveness, we used the net monetary benefit (NMB), which was calculated as the difference between two measures: 1) the total effectiveness times the willingness-to-pay (WTP) threshold, which values the effectiveness generated by the intervention in terms of the opportunity cost forgone and 2) the total cost of the intervention. We set the WTP threshold to US $95,958, according to the latest comprehensive report, which calculated this value for 174 countries.^24^ We evaluated the following clinical outcomes, i.e., the probabilities of reoperation due to missed MGDs, BNE, persistent postoperative hypoparathyroidism, and persistent RLN injury, by calculating the proportion of people experiencing these events in the simulated cohort. To simplify the model, we did not account for radiation exposure for preoperative imaging modalities (4D-CT, SPECT/CT, and FCH-PET/CT).

### 2.3 Sensitivity and uncertainty analyses

To determine the influence of each model parameter on the model outcomes, we conducted a one-way sensitivity analysis by varying the parameter values within ±50% of their base case values and evaluating the resulting effect on the NMBs. The sensitivity analysis was performed on all parameters, and its result was presented as a tornado diagram, which ranks model parameters according to their influences. To account for the effect of uncertainty in the model parameter estimates, we conducted an uncertainty analysis using a Monte Carlo sampling approach. For probabilities and utility weights, we fitted beta distributions, where the parameters were estimated by using the method of moments.^25^ For costs, gamma distributions were used, and we followed the same approach for estimating their parameters. When 95% confidence intervals (CI) were available, we assumed that the CIs were 1.98*2 standard deviations wide. When only the lower and upper limits were reported, we assumed the limits to be equal to the 95% CIs. We generated 5,000 random samples of the parameters’ estimates from the corresponding beta and gamma distributions and evaluated our model at each sample. We presented the results of the uncertainty analysis using scatter plots and cost-effectiveness acceptability curves. To identify the values of the influential parameters at which the conclusions of the cost-effectiveness analyses change (from being cost-effective to not cost-effective), we performed threshold analyses. The selection of the influential parameters was based on the results of the sensitivity analyses.

The construction of the decision-tree model and the analyses were performed using Python 3.9. The Python codes are publicly available at https://github.com/danielbatora/batora_phpt_cea.

## 3. RESULTS

### 3.1 Cost-effectiveness analysis

#### 3.1.1 Base-case analysis

The least expensive preoperative localization modality was 4D-CT with $10,289 and 13.93 QALYs, which was followed by ultrasound with $10,740 and 13.89 QALYs and the SPECT/CT with $10,778 and 13.91 QALYs. FCH-PET/CT was the most expensive modality with a cost of $11,623, albeit it provided 13.94 QALYs (S1 Table). At a $95,958 WTP threshold, FCH-PET/CT resulted in a NMB value of $264. For all imaging modalities, the addition of ioPTH monitoring generated higher costs and fewer QALYs, thereby making them dominated strategies (Table 2). The use of the Miami criterion was found to be less expensive and associated with more QALYs in all cases when compared to using the Vienna criterion (Table 2).

**Table 2.** Base case analysis for all interventions.

| Intervention | Total costs (\$) | QALYs | Incremental costs (\$) | Incremental QALYs | INMB |
| --- | --- | --- | --- | --- | --- |
| 4D-CT without ioPTH monitoring | 10289 | 13,926 | 0 | 0 | 0 |
| Ultrasound without ioPTH monitoring | 10740 | 13,893 | 451 | -0,03 | -3585 |
| SPECT/CT without ioPTH monitoring | 10778 | 13,913 | 37 | 0,02 | 1839 |
| 4D-CT with ioPTH monitoring (Miami criterion) | 10808 | 13,925 | 31 | 0,01 | 1122 |
| 4D-CT with ioPTH monitoring (Vienna criterion) | 10971 | 13,915 | 163 | -0,01 | -1128 |
| Ultrasound with ioPTH monitoring (Miami criterion) | 11286 | 13,892 | 315 | -0,02 | -2440 |
| Ultrasound with ioPTH monitoring (Vienna criterion) | 11371 | 13,911 | 85 | 0,02 | 1689 |
| SPECT/CT with ioPTH monitoring (Miami criterion) | 11400 | 13,886 | 29 | -0,03 | -2461 |
| SPECT/CT with ioPTH monitoring (Vienna criterion) | 11535 | 13,902 | 134 | 0,02 | 1430 |
| FCH-PET/CT without ioPTH monitoring | 11623 | 13,942 | 88 | 0,04 | 3797 |
| FCH-PET/CT with ioPTH monitoring (Miami criterion) | 12227 | 13,940 | 604 | 0,00 | -838 |
| FCH-PET/CT with ioPTH monitoring (Vienna criterion) | 12449 | 13,928 | 222 | -0,01 | -1394 |

*Abbreviations: FCH-PET/CT, <sup>18</sup>F-fluorocholine positron emission tomography; INMB,* *incremental net monetary benefit; ioPTH, intraoperative parathyroid hormone; QALYs, quality-* *adjusted life years; SPECT/CT, <sup>99m</sup>Tc-Sestamibi single-photon-emission computed* *tomography/computed tomography; 4D-CT, four-dimensional-CT.*

#### 3.1.2 Sensitivity analysis

We conducted two one-way sensitivity analyses focusing on comparing the non-dominated preoperative localization strategies, and ioPTH monitoring. In the comparison between the non-dominated preoperative localization strategies without ioPTH monitoring, i.e., the 4D-CT compared to FCH-PET/CT, the model was most sensitive to the prevalence of single adenomas with NMB ranging from -$6,909 to $2,698, the sensitivity of FCH-PET/CT for single adenomas with NBM ranging from -$6,651 to $1716, the cost of the FCH-PET/CT with NMB ranging from -$726 to $1254 and the PPV of FCH-PET/CT for single adenomas and MGD with NMB ranging from -$4,115 to $488 and -$2,080 to $335, respectively (Fig 2A).

**Figure 2.**
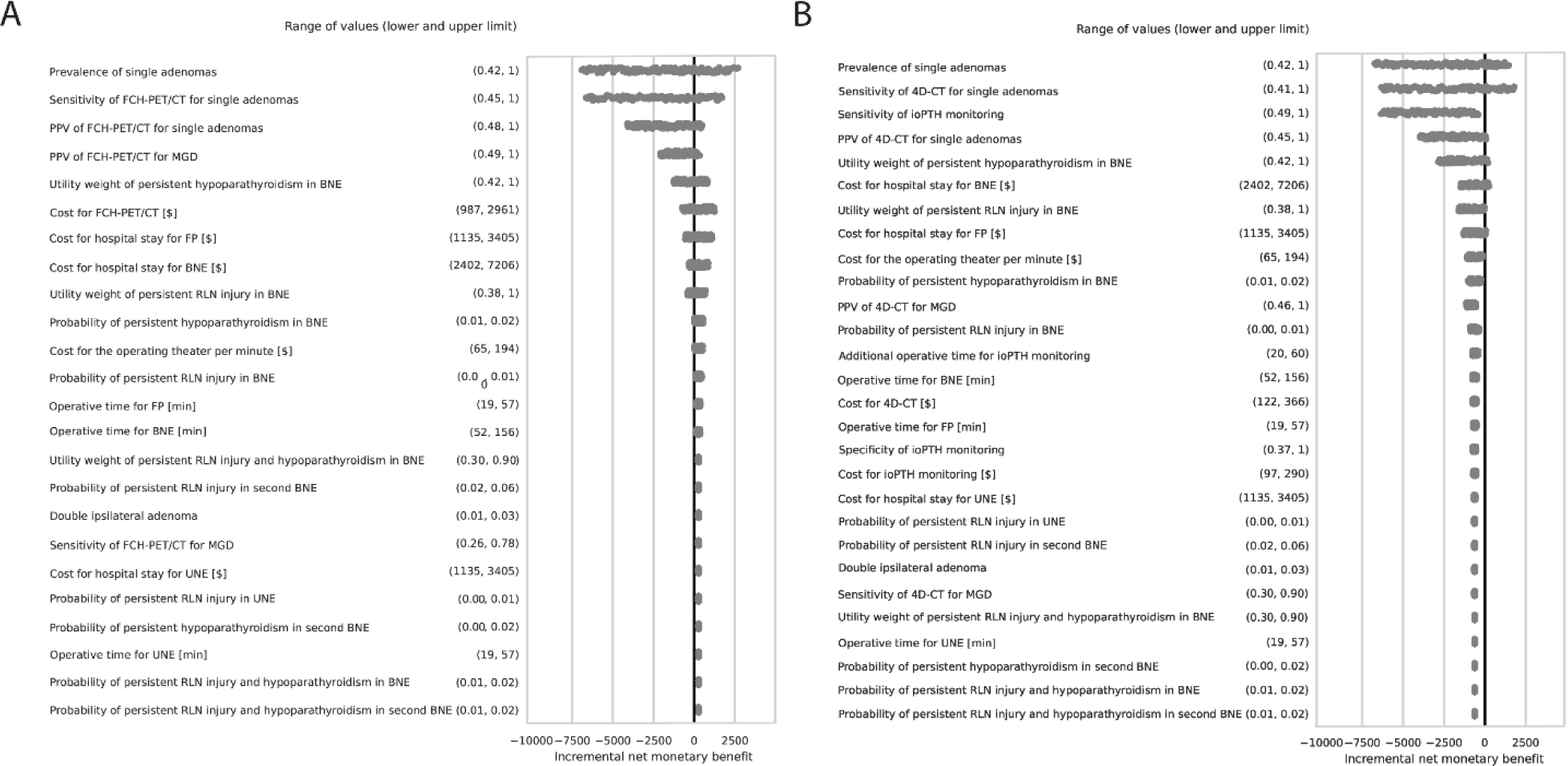
One-way sensitivity analysis. **A:** Variation of the parameters of the ^18^F-fluorocholine positron emission tomography (FCH-PET/CT) without intraoperative parathyroid hormone (ioPTH) monitoring within ±50% of their base case values. Incremental net monetary benefits were calculated in comparison with the base case values of the four-dimensional-computed tomography (4D-CT) without ioPTH monitoring, as this was the least expensive intervention in our base case analysis. **B:** Varying the parameters were done in a similar manner as in panel A. Incremental net monetary benefits of 4D-CT with ioPTH monitoring were calculated in comparison with the base case values of the 4D-CT without ioPTH monitoring. *Abbreviations: BNE, bilateral neck exploration; FP, focused parathyroidectomy; MGD, multiglandular disease; PPV, positive predictive value; RLN, recurrent laryngeal nerve; UNE, unilateral neck exploration*.

In the comparison of the with and without ioPTH monitoring strategies for 4D-CT imaging, the model was most sensitive to the prevalence of solitary adenomas with NMB ranging from -$6,743 to $1,452, followed by the sensitivity of the 4D-CT for single adenomas with NMB ranging from -$6,332 to $1,800, the sensitivity of ioPTH monitoring with NMB ranging from -$6,349 to -$409, and the PPV of 4D-CT for single adenomas with NMB ranging from -$6,349 to -$409. Furthermore, the results were sensitive to the cost for outpatient and inpatient hospital stay (-$1,473 – $242, -$1,306 – $71), the utility weights for complications (persistent hypoparathyroidism [-$2,849 – $192] and RLN injury [-$1,590 – -$30]) and the cost of the operating theater per minute (-$1,150 – -$238) (Fig 2B).

#### 3.1.3 Uncertainty analysis

When accounting for uncertainties in all parameters, the 4D-CT without ioPTH monitoring was the least expensive strategy with $10,302 (95% CI $10,297-$10,307) and 13.926 QALYs (95% CI 13.942–13.943). The most expensive and the only non-dominated intervention was the FCH-PET/CT with $11,622 (95% CI $11,618–$11,627) and 13.943 QALYs (95% CI 13.942–13.943) (S2 Table). Varying the WTP thresholds from $20,000 to $500,000 revealed that the FCH-PET/CT is unlikely to be cost-effective at WTP thresholds below $80,000 (Fig 3).

**Figure 3.**
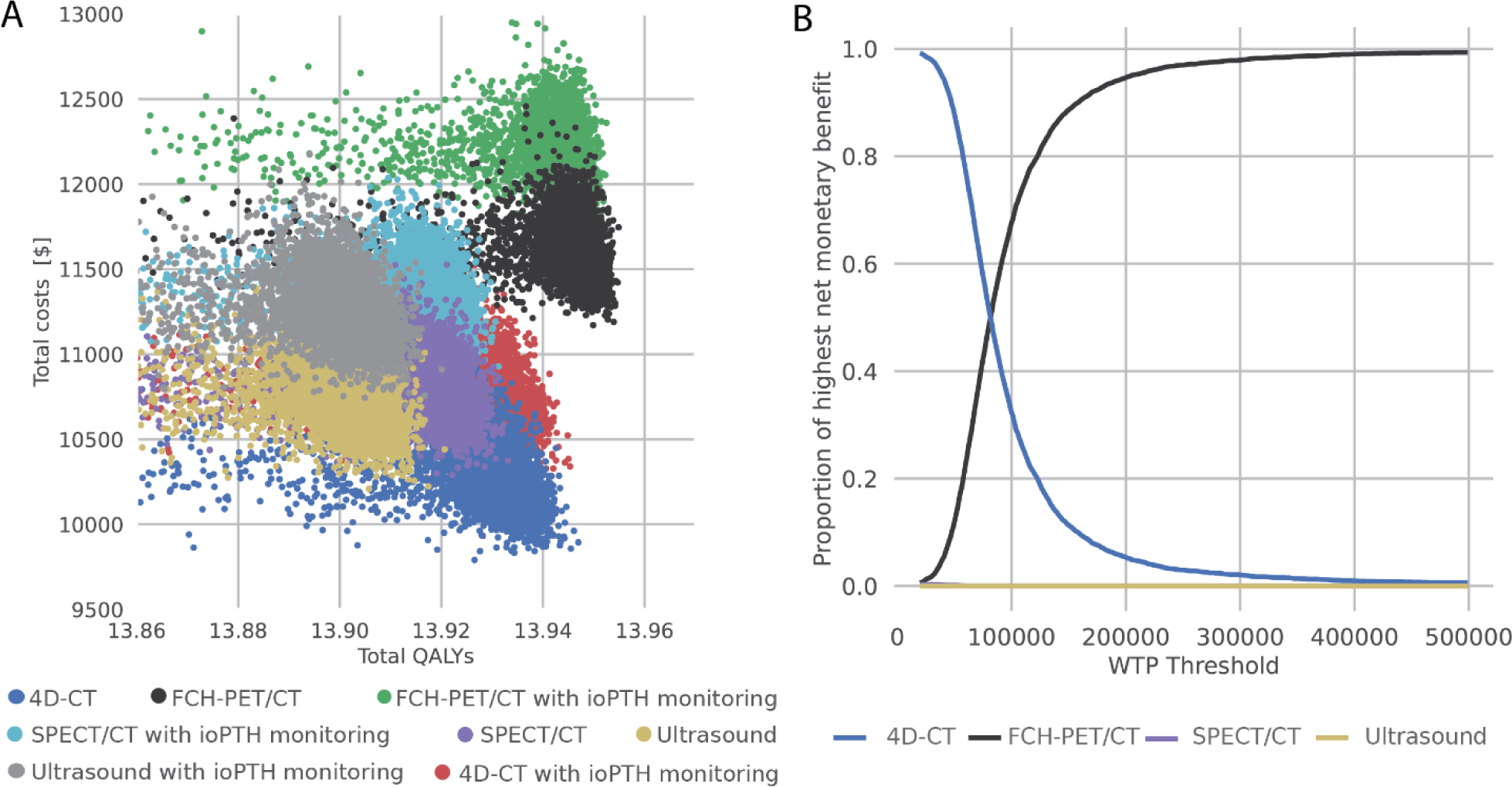
Uncertainty analysis. **A:** Scatter plots showing the outcome of the Monte Carlo simulation. For each parameter, beta or gamma distributions were fitted. In all, 5,000 samples were drawn from the distributions and were used for the subsequent analysis in the decision-tree model. Total costs and quality-adjusted life years (QALYs) were calculated for each sample. **B:** A cost-effectiveness acceptability curve was generated from the result of the Monte Carlo simulation. We varied the willingness-to-pay (WTP) threshold from 20,000 to 500,000; and calculated the proportion of the highest net monetary benefit (NMB) for each intervention. Below a WTP threshold of $80,000, 4D-CT without intraoperative parathyroid hormone (ioPTH) monitoring yields the biggest proportion of highest NMBs. Above a WTP threshold of $80,000, ^18^F-fluorocholine positron emission tomography (FCH-PET/CT) becomes the dominant strategy. In the range of a WTP threshold between $20,000 and $500,000, ^99m^Tc-Sestamibi single-photon-emission computed tomography [SPECT]/computed tomography [CT] and ultrasound are dominated strategies. *Abbreviation: 4D-CT, four-dimensional-CT*.

#### 3.1.4 Threshold analysis

For 4D-CT, we identified the values for different parameters that would render the ioPTH monitoring with the Miami criterion cost-effective: an MGD prevalence of more than 49%, a positive predictive value of 4D-CT for detecting MGD of less than 70%, a probability of persistent hypoparathyroidism due to reoperation of more than 39%, and a probability of persistent RLN injury due to reoperation of more than 28% (Fig S1).

### 3.2 Clinical outcomes by using the Miami or Vienna criterion for ioPTH monitoring

By using 4D-CT with the Miami criterion, the rate of reoperations decreased from 10.50 to 2.77 per 1,000 patients compared to not using ioPTH monitoring. With the Vienna criterion, which has a higher specificity, the rate of reoperations decreased to 0.57 per 1,000 patients compared to not using ioPTH monitoring.

With the Miami criterion, the rate of BNEs increased from 257.48 to 274.38 per 1,000 patients, while it increased to 347.45 per 1,000 patients with the Vienna criterion due to its lower sensitivity compared to not using ioPTH monitoring.

The rate of complications associated with the surgery was higher with the Vienna criterion: the overall probability of persistent hypoparathyroidism and persistent RLN injury increased from 2.91 to 3.81 per 1,000 patients and from 1.68 to 1.74 per 1,000 patients, respectively, compared to not using ioPTH monitoring. With the Miami criterion, the rate of persistent hypoparathyroidism increased to 3.03 per 1,000 patients, whereas the rate of persistent RLN injury decreased to 1.45 per 1,000 patients compared to not using ioPTH monitoring (Fig 4).With all the other preoperative imaging modalities, the trends were generally similar (S3 Table).

**Figure 4.**
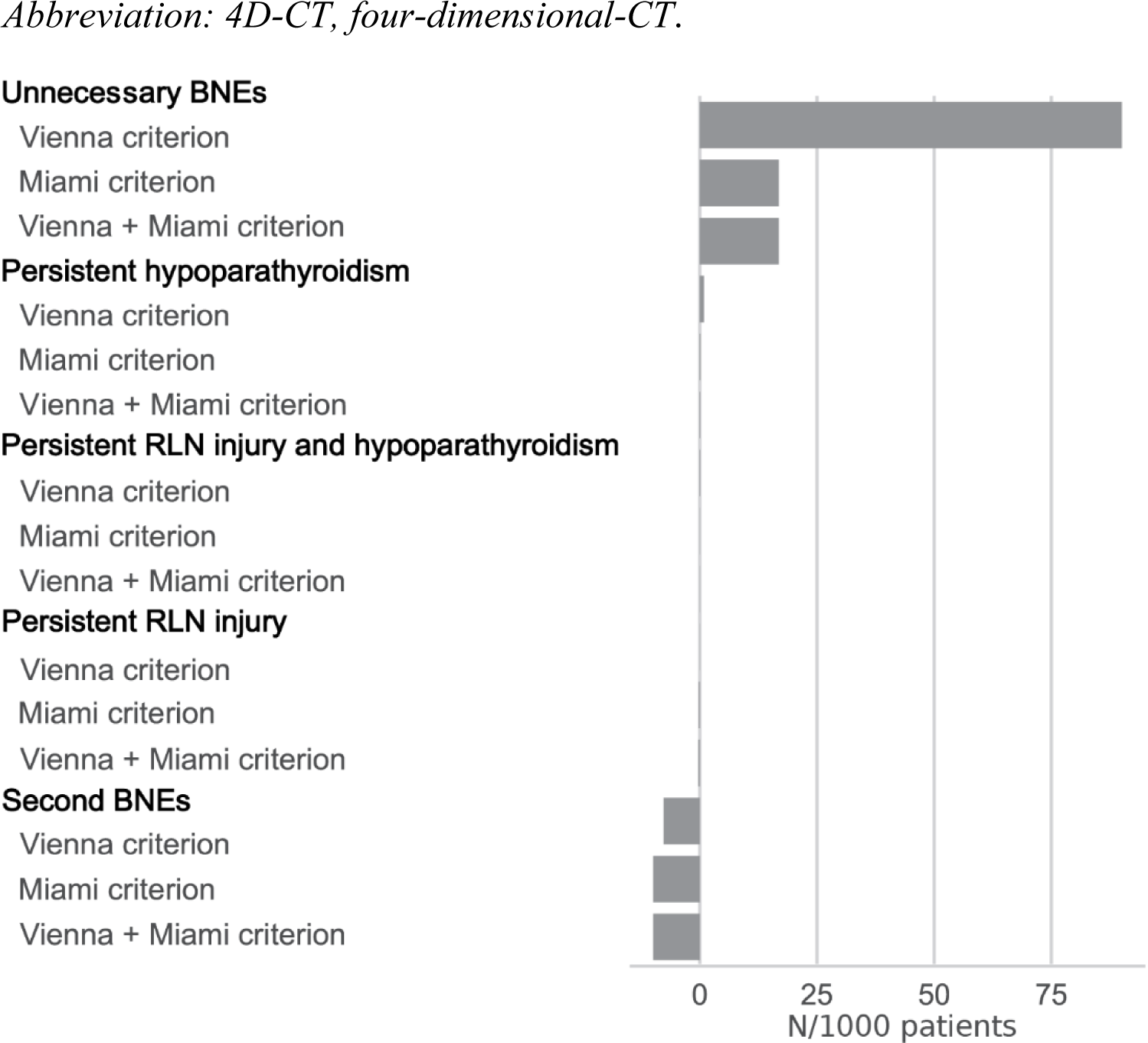
The impact of different ioPTH criteria on the main clinical events using four-dimensional-computed tomography (4D-CT) Differences in clinical events per 1,000 parathyroidectomies compared with patients without intraoperative parathyroid hormone (ioPTH) monitoring. In addition to the Miami and Vienna criterion, we evaluated the combined use of the Miami criterion for solitary adenomas and the Vienna criterion for suspected multiglandular disease. Unnecessary bilateral neck explorations (BNEs) were minimized by applying the Miami criterion (+17 BNEs compared to patients without ioPTH monitoring), while reoperations were minimized by using the Vienna criterion (−10 reoperations compared to patients without ioPTH monitoring). *Abbreviation: RLN, recurrent laryngeal nerve*.

## 4. DISCUSSION

This study evaluated the clinical impact and cost-effectiveness of pre- and intraoperative diagnostic tools in the surgical management of PHPT in the United States. The study has three major results. First, 4D-CT alone was found to be the least extensive preoperative localization strategy and FCH-PET/CT to be cost-effective near the US WTP threshold. Second, the addition of ioPTH monitoring decreased the frequency of reoperations; however, it was not cost-effective due to a higher rate of BNEs, in addition to the assay and anesthesia costs. Third, the rate of BNEs was considerably higher by using the Vienna criterion compared to the Miami criterion.

An accurate localization of parathyroid adenoma(s) is an important component of the preoperative planning of parathyroid surgery. Our analysis demonstrated that 4D-CT was the least expensive preoperative imaging modality, while both SPECT/CT and ultrasound were more expensive and provided fewer QALYs than 4D-CT. FCH-PET/CT was superior to 4D-CT in terms of the QALYs gained and is likely cost-effective below a $85,000 WTP threshold. Our results were consistent with the most recent cost-effectiveness analysis^14^ despite using more conservative estimates for the diagnostic performance parameters for several preoperative localization strategies (ultrasound, Sestamibi-SPECT/CT, and 4D-CT) from the latest meta-analyses. A plausible explanation for reaching a comparable conclusion might lie in the inclusion of MGD cases in our study, for which the preoperative localization is considerably less accurate.^6,7^

Even though our analysis showed 4D-CT imaging to be the least expensive strategy, the use of 4D-CT as the first-in-line preoperative imaging modality in the future could be restricted to centers with a high frequency of PHPT cases, as its operation requires considerable radiological training. Furthermore, due to limited available data, our model could not evaluate the cost-effectiveness of the sequential application of preoperative imaging modalities, which is commonly applied in clinical practice to reserve the more expensive modalities for cases where the first-in-line treatment is likely less accurate.

In our analysis, the addition of ioPTH monitoring was not cost-effective, even though it led to a significantly reduced number of reoperations. The clinical benefit of ioPTH monitoring was outweighed by erroneous classifications by the method, resulting in a substantial increase of unnecessary BNEs. Additionally, the sensitivity analysis revealed that the sensitivity of the assay and the occupancy of the operating theater were key determinants in the cost-effectiveness of ioPTH monitoring. In line with our results, the study of Badii et al. showed that ioPTH monitoring was not cost-saving due to substantially longer operative times.^13^ Morris et al. suggested that the use of ioPTH monitoring would only be beneficial in a relatively small fraction of patients where the accuracy of preoperative localization was low.^12^ This finding is supported by our threshold analysis, which revealed that only a PPV for MGD of less than 70% would render ioPTH monitoring cost-effective. Recent technological innovations in the field aim to substantially reduce both cost and time expenditure for ioPTH monitoring, which suggests the possibility of being cost-effective in the future.^26,27^

Overall, the interpretation of our cost-effectiveness analysis is limited to the United States. However, our uncertainty analysis suggested that the decision-analytical model is robust and, therefore, can be adapted to other countries with distinct cost structures as well.

In addition to the cost-effectiveness, reducing the complications in parathyroid surgery are essential. We found that ioPTH monitoring using the Vienna criterion minimized the risk of reoperations, while the Miami criterion minimized complication rates. The physical manipulation of an adenoma during operation might lead to an increase in ioPTH.^3^ This risk could be higher in the case of MGD due to the manipulation of more than one gland. The Miami criterion has a higher risk of false positive results due to using the highest ioPTH level (pre-incision or pre-excision) as the baseline value. Surgeons should therefore consider using the Vienna criterion for patients with suspected MGD to maximize specificity and the Miami criterion for patients with suspected solitary adenoma to maximize sensitivity.

### 4.1 Limitations of the study

As the use of FCH-PET/CT and 4D-CT imaging has only been recently introduced for the localization of parathyroid adenomas, there were only a handful of available systematic reviews to inform the estimates of the model parameters related to the imaging modality. Hence, for the sensitivity and PPV values for FCH-PET/CT in MGD, we relied on data from a single institutional report^9^, which limits the generalizability of our results. Similarly, data for estimating the model parameters related to the ioPTH protocol were derived from a limited number of studies, which showed considerable variations in the estimates. To overcome this limitation in the evidence base, we conducted sensitivity analyses to determine how robust the conclusions were to variation in the model parameter values. Furthermore, our uncertainty analysis accounted for the effect of uncertainty in the estimates of the model parameters on the cost-effectiveness analysis results. Nevertheless, meta-analyses that stratify solitary adenomas and MGD are warranted to reduce the risk of bias in the performance values of localization techniques.

Moreover, the true rate of solitary adenomas is also debated, as some institutions report a significantly higher rate of MGD than the consensus value.^2^ Accordingly, our threshold analysis reveals that the prevalence of MGD strongly impacts the complication rates which influences the cost-effectiveness of ioPTH monitoring. Therefore, it is critical to consider the existing institutional variations in this parameter during the decision-making process. In addition, as FCH-PET/CT is not yet authorized in the US, our model likely underestimates the future cost of FCH by inferring from the cost of the most used PET tracer, ^18^F-fluorodeoxyglucose (CPT A9552); suggesting that the threshold to reach cost-effectiveness for FCH-PET/CT might be higher than estimated. Similarly, as a societal perspective was not incorporated into our evaluation, e.g., absence from work due to inpatient hospitalization, the economic consequences of unnecessary BNEs due to ioPTH monitoring might be underestimated in our model.

### 4.2 Conclusions

In our decision-analytical model of parathyroidectomy that also considered the MGD cases, 4D-CT was found to be the least expensive diagnostic tool for the preoperative localization of parathyroid adenomas, and FCH-PET/CT is likely a cost-effective modality. Due to an excessive increase of BNEs, the use of ioPTH monitoring was found not cost-effective in PHPT but led to a significant reduction in the rate of reoperations. If ioPTH monitoring is used, our model suggest that the Miami criterion should be applied for suspected solitary adenomas and the Vienna criterion for suspected MGD.

## Data Availability

All data produced are publicly available at https://github.com/danielbatora/batora_phpt_cea.

https://github.com/danielbatora/batora_phpt_ce

## Acknowledgements

We would like to thank Prof. Dr. med. Thomas J. Musholt, Section of Endocrine Surgery, Department of General, Visceral and Transplantation Surgery, University Medical Center, Johannes Gutenberg University Mainz, Germany for his critical revision of the manuscript and the valuable inputs.

---

**Figure S1.**
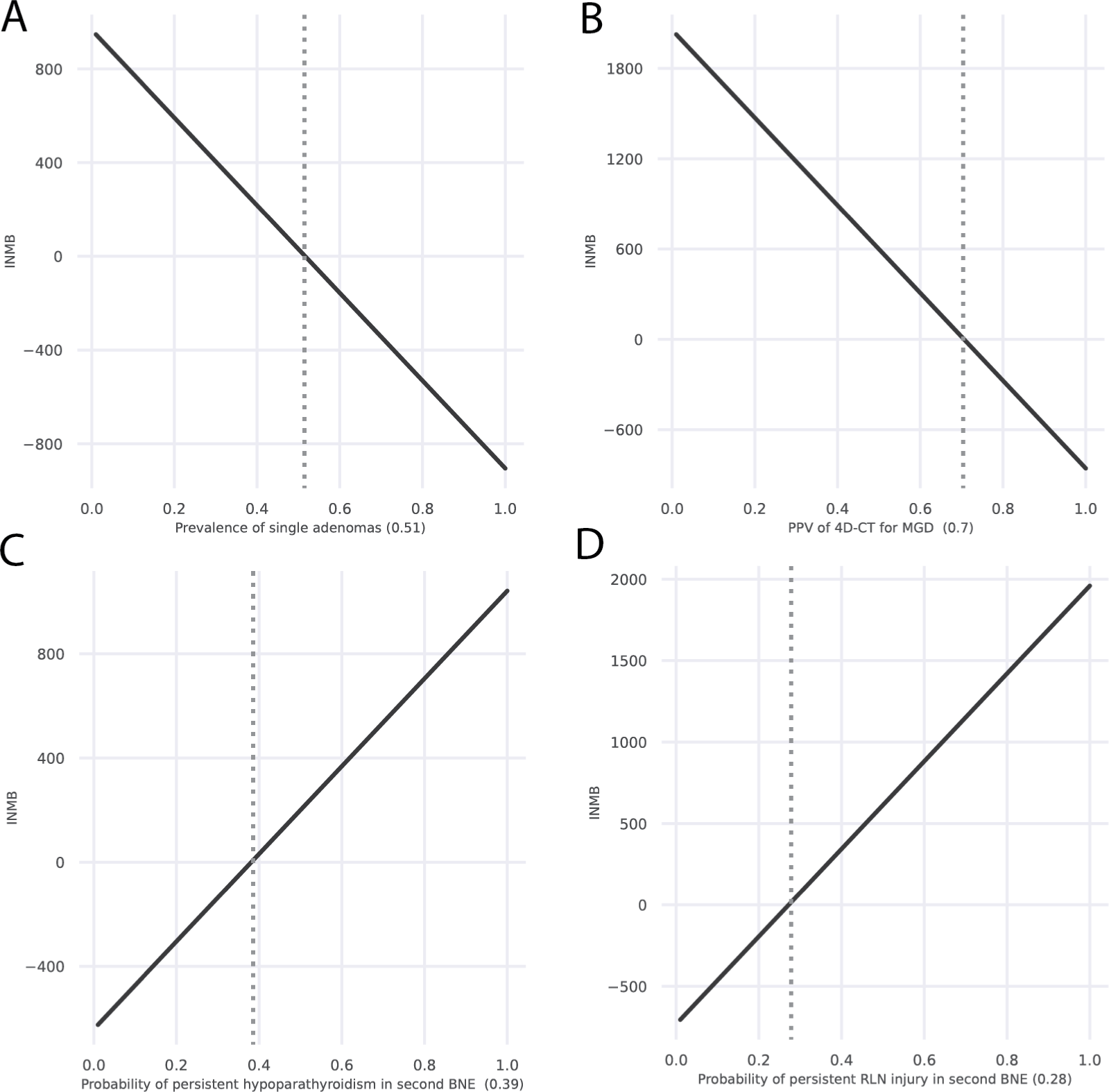
Threshold analysis on parameters for the four-dimensional-CT (4D-CT). Assessment of the theoretical possibility of intraoperative parathyroid hormone (ioPTH) monitoring with a positive net monetary benefit value. Each parameter of the 4D-CT with ioPTH monitoring (Miami criterion) was varied from 0 to 1 for probabilities and utility weights, respectively, and 0 to the base case value for costs. Incremental net monetary benefit (INMB) was calculated in comparison with the 4D-CT without ioPTH monitoring. *Abbreviations: BNE, bilateral neck exploration; MGD, multiglandular disease; PPV, positive predictive value; RLN, recurrent laryngeal nerve; 4D-CT, four-dimensional-CT*.

**S1 Table.** Total costs and quality-adjusted life years (QALYs) for the non-dominated interventions of the base case analysis.

| Intervention | Total costs [\$] | Total QALYs | Incremental costs [\$] | Incremental QALYs | INMB |
| --- | --- | --- | --- | --- | --- |
| 4D-CT without ioPTH | 10,289 | 13.926 | 0 | 0.00 | 0 |
| FCH-PET/CT without ioPTH | 11,623 | 13.942 | 1334 | 0.02 | 264 |

*Abbreviation: INMB, incremental net monetary benefits.*

**S2 Table.**
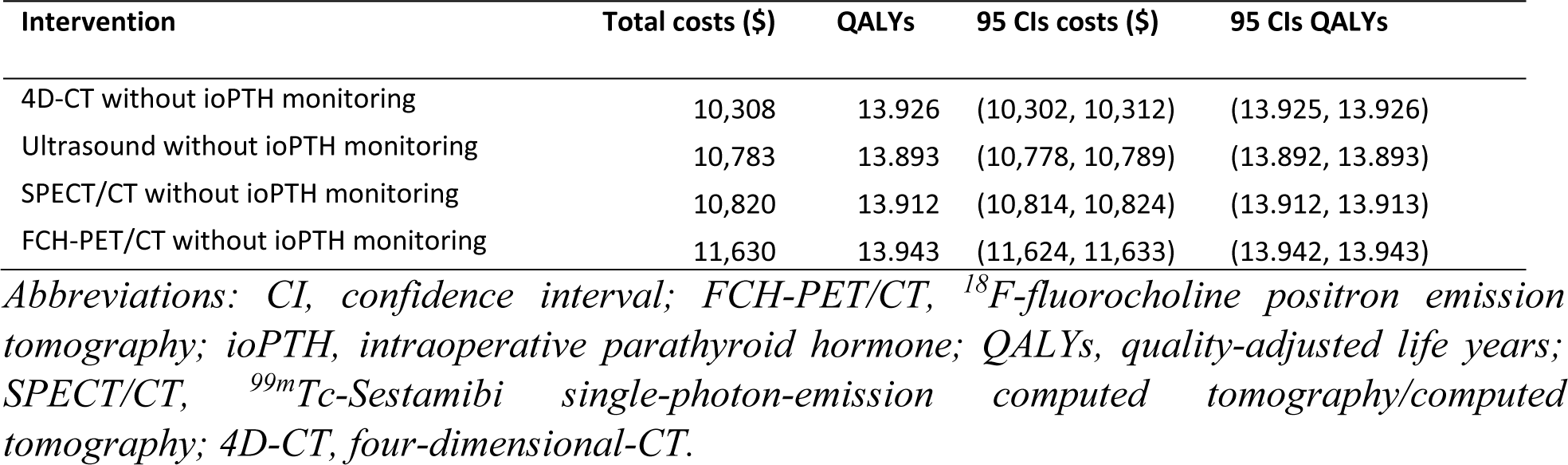
Confidence intervals of the monte-carlo simulation.

| Intervention | Total costs (\$) | QALYs | 95 CIs costs (\$) | 95 CIs QALYs |
| --- | --- | --- | --- | --- |
| 4D-CT without ioPTH monitoring | 10,308 | 13.926 | (10,302, 10,312) | (13.925, 13.926) |
| Ultrasound without ioPTH monitoring | 10,783 | 13.893 | (10,778, 10,789) | (13.892, 13.893) |
| SPECT/CT without ioPTH monitoring | 10,820 | 13.912 | (10,814, 10,824) | (13.912, 13.913) |
| FCH-PET/CT without ioPTH monitoring | 11,630 | 13.943 | (11,624, 11,633) | (13.942, 13.943) |

*Abbreviations: CI, confidence interval; FCH-PET/CT, <sup>18</sup>F-fluorocholine positron emission* *tomography; ioPTH, intraoperative parathyroid hormone; QALYs, quality-adjusted life years;* *SPECT/CT, <sup>99m</sup>Tc-Sestamibi single-photon-emission computed tomography/computed* *tomography; 4D-CT, four-dimensional-CT.*

**S3 Table.** Number of clinical events per 1000 patients for all interventions.

| Intervention | BNE | Reoperations | Persistent hypoparathyroidism | Persistent RLN injury | Persistent hypoparathyroidism and RLN injury |
| --- | --- | --- | --- | --- | --- |
| 4D-CT without ioPTH monitoring | 257.45 | 10.50 | 2.91 | 1.68 | 0.02 |
| 4D-CT with ioPTH monitoring (Miami criterion) | 274.34 | 2.77 | 3.03 | 1.45 | 0.02 |
| 4D-CT with ioPTH monitoring (Vienna criterion) | 347.45 | 0.58 | 3.81 | 1.74 | 0.02 |
| FCH-PET/CT without ioPTH monitoring | 239.68 | 3.00 | 2.62 | 1.30 | 0.05 |
| FCH-PET/CT with ioPTH monitoring (Miami criterion) | 257.29 | 0.78 | 2.82 | 1.30 | 0.02 |
| FCH-PET/CT with ioPTH monitoring (Vienna criterion) | 333.98 | 0.17 | 3.66 | 1.66 | 0.02 |
| SPECT/CT without ioPTH monitoring | 284.55 | 6.00 | 3.10 | 1.64 | 0.08 |
| SPECT/CT with ioPTH monitoring (Miami criterion) | 300.84 | 1.56 | 3.29 | 1.55 | 0.03 |
| SPECT/CT with ioPTH monitoring (Vienna criterion) | 371.52 | 0.33 | 4.07 | 1.85 | 0.02 |
| Ultrasound without ioPTH monitoring | 371.14 | 3 | 4.06 | 1.95 | 0.05 |
| Ultrasound with ioPTH monitoring (Miami criterion) | 385.59 | 0.79 | 4.22 | 1.94 | 0.03 |
| Ultrasound with ioPTH monitoring (Vienna criterion) | 448.31 | 0.16 | 4.91 | 2.22 | 0.03 |

*Abbreviations: BNE, bilateral neck exploration; FCH-PET/CT, <sup>18</sup>F-fluorocholine positron* *emission tomography; ioPTH, intraoperative parathyroid hormone; RLN, recurrent laryngeal* *nerve; SPECT/CT,*
*<sup>99m</sup>Tc-Sestamibi single-photon-emission computed tomography/computed tomography; 4D-CT,* *four-dimensional-CT.*

## LIST OF ABBREVIATIONS

4D: four-dimensional
BNE: bilateral neck exploration
CF: conversion factor
CI: confidence interval
CPT: Current Procedural Terminology
CT: computed tomography
DRG: Diagnostic-Related Group
FCH-PET: ^18^F-fluorocholine positron emission tomography
FP: focused parathyroidectomy
ioPTH: intraoperative parathyroid hormone
MGD: multiglandular disease
NIH: National Institutes of Health
NMB: net monetary benefit
PHPT: primary hyperparathyroidism
RLN: recurrent laryngeal nerve
SPECT: single-photon-emission computed tomography
UNE: unilateral neck exploration
WTP: willingness-to-pay

## References

1. Yeh MW, Ituarte PH, Zhou HC, Nishimoto S, Liu IL, Harari A, et al. Incidence and prevalence of primary hyperparathyroidism in a racially mixed population. The Journal of clinical endocrinology and metabolism. 2013(98(3)):1122-1129.

2. Fraker DL, Harsono H, Lewis R. Minimally Invasive Parathyroidectomy: Benefits and Requirements of Localization, Diagnosis, and Intraoperative PTH Monitoring. Long-Term Results. World J Surg. 2009;33(11):2256–2265.

3. Khan AA, Hanley DA, Rizzoli R, et al. Primary hyperparathyroidism: review and recommendations on evaluation, diagnosis, and management. A Canadian and international consensus. Osteoporosis international: a journal established as result of cooperation between the European Foundation for Osteoporosis and the National Osteoporosis Foundation of the USA. 2017;28(1):1–19.

4. Grant CS, Thompson G, Farley D, van Heerden J. Primary hyperparathyroidism surgical management since the introduction of minimally invasive parathyroidectomy: Mayo Clinic experience. Archives of surgery (Chicago, Ill.: 1960). 2005;140(5):472–8; discussion 478-9.

5. El-Hady HA, Radwan HS. Focused parathyroidectomy for single parathyroid adenoma: a clinical account of 20 patients. Electronic physician. 2018;10(6):6974–6980.

6. Patel DD, Bhattacharjee S, Pandey AK, Kopp CR, Ashwathanarayana AG, Patel HV, et al. Comparison of 4D computed tomography and F-18 fluorocholine PET for localisation of parathyroid lesions in primary hyperparathyroidism: A systematic review and meta-analysis. Clinical endocrinology. 2023(99(3)):262-271.

7. Ruda JM, Hollenbeak CS, Stack BC. A systematic review of the diagnosis and treatment of primary hyperparathyroidism from 1995 to 2003. Otolaryngol Head Neck Surg. 1995(132(3):):359-372.

8. Kattar N, Migneron M, Debakey MS, Haidari M, Pou AM, McCoul ED. Advanced Computed Tomographic Localization Techniques for Primary Hyperparathyroidism: A Systematic Review and Meta-analysis. JAMA otolaryngology--head & neck surgery. 2022;148(5):448-456.

9. Talbot J-N, Périé S, Tassart M, et al. 18F-fluorocholine PET/CT detects parathyroid gland hyperplasia as well as adenoma: 401 PET/CTs in one center. The quarterly journal of nuclear medicine and molecular imaging: official publication of the Italian Association of Nuclear Medicine (AIMN) [and] the International Association of Radiopharmacology (IAR), [and] Section of the Society of… 2023;67(2):96–113.

10. Bilezikian JP, Khan AA, Silverberg SJ, et al. Evaluation and Management of Primary Hyperparathyroidism: Summary Statement and Guidelines from the Fifth International Workshop. Journal of bone and mineral research: the official journal of the American Society for Bone and Mineral Research. 2022;37(11):2293–2314.

11. Barczynski M, Konturek A, Hubalewska-Dydejczyk A, Cichon S, Nowak W. Evaluation of Halle, Miami, Rome, and Vienna intraoperative iPTH assay criteria in guiding minimally invasive parathyroidectomy. Langenbeck’s archives of surgery. 2009;394(5):843–849.

12. Morris LF, Zanocco K, Ituarte PHG, et al. The value of intraoperative parathyroid hormone monitoring in localized primary hyperparathyroidism: a cost analysis. Annals of surgical oncology. 2010;17(3):679–685.

13. Badii B, Staderini F, Foppa C, et al. Cost-benefit analysis of the intraoperative parathyroid hormone assay in primary hyperparathyroidism. Head & neck. 2017;39(2):241–246.

14. Yap A, Hope TA, Graves CE, et al. A cost-utility analysis of 18F-fluorocholine-positron emission tomography imaging for localizing primary hyperparathyroidism in the United States. Surgery. 2022;171(1):55–62.

15. Arias E XJ. United States Life Tables, 2019. 2022;Natl Vital Stat Rep(70(19)):1-59.

16. Zanocco K, Sturgeon C. How should age at diagnosis impact treatment strategy in asymptomatic primary hyperparathyroidism? A cost-effectiveness analysis. Surgery. 2008;144(2):290–298.

17. Wilhelm SM, Wang TS, Ruan DT, et al. The American Association of Endocrine Surgeons Guidelines for Definitive Management of Primary Hyperparathyroidism. JAMA surgery. 2016;151(10):959–968.

18. Riss P, Kaczirek K, Heinz G, Bieglmayer C, Niederle B. A “defined baseline” in PTH monitoring increases surgical success in patients with multiple gland disease. Surgery. 2007;142(3):398–404.

19. Weinstein MC. Recent developments in decision-analytic modelling for economic evaluation. PharmacoEconomics. 2006;24(11):1043–1053.

20. Medican Reimbursement Schedule. Available at: https://www.medicare.gov/procedure-price-lookup. Accessed October 01, 2023. 2023. Available at: www.medicare.gov/procedure-price-lookup.

21. Anesthesiologists Center | CMS. 2022. Available at: https://www.cms.gov/Center/Provider-Type/Anesthesiologists-Center. Accessed October 01, 2023. 2022. Available at: www.cms.gov/Center/Provider-Type/Anesthesiologists-Center.

22. Lubitz CC, Stephen AE, Hodin RA, Pandharipande P. Preoperative localization strategies for primary hyperparathyroidism: an economic analysis. Annals of surgical oncology. 2012;19(13):4202–4209.

23. Tengs TO, Wallace A. One thousand health-related quality-of-life estimates. Medical care. 2000;38(6):583–637.

24. Pichon-Riviere A, Drummond M, Palacios A, Garcia-Marti S, Augustovski F. Determining the efficiency path to universal health coverage: cost-effectiveness thresholds for 174 countries based on growth in life expectancy and health expenditures. The Lancet. Global health. 2023(11(6)):e833–e842.

25. Briggs AH, Goeree R, Blackhouse G, O’Brien BJ. Probabilistic analysis of cost-effectiveness models: choosing between treatment strategies for gastroesophageal reflux disease. Medical decision making: an international journal of the Society for Medical Decision Making. 2002;22(4):290–308.

26. Jarrige V, Nieuwenhuis JH, van Son JPHF, Martens MFWC, Vissers JLM. A fast intraoperative PTH point-of-care assay on the Philips handheld magnotech system. Langenbeck’s archives of surgery. 2011;396(3):337–343.

27. Kurzawinski T, Soromani C, Aziz TA, Lam F, Garcia RV, Matias M, et al. New, simple, fast, whole blood Intraoperative PTH assay – Laboratory and Clinical validation. Br J Surg. 2022(109.).

28. Kebebew E, Duh QY, Clark OH. Total thyroidectomy or thyroid lobectomy in patients with low-risk differentiated thyroid cancer: surgical decision analysis of a controversy using a mathematical model. World journal of surgery. 2000;24(11):1295–1302.

29. Spector BC, Netterville JL, Billante C, Clary J, Reinisch L, Smith TL. Quality-of-life assessment in patients with unilateral vocal cord paralysis. Otolaryngology--head and neck surgery: official journal of American Academy of Otolaryngology-Head and Neck Surgery. 2001;125(3):176–182.

